# A central serotonin regulating gene polymorphism (TPH2) determines vulnerability to acute tryptophan depletion-induced anxiety and ventromedial prefrontal threat reactivity

**DOI:** 10.1101/2023.03.18.23287402

**Authors:** Congcong Liu, Keshuang Li, Meina Fu, YingYing Zhang, Cornelia Sindermann, Christian Montag, Xiaoxiao Zheng, Hongxing Zhang, Yao Shuxia, Zheng Wang, Bo Zhou, Keith M. Kendrick, Benjamin Becker

**Affiliations:** The Center of Psychosomatic Medicine, Sichuan Provincial Center for Mental Health, Provincial People’s Hospital, University of Electronic Science and Technology of China, Chengdu, China; School of Psychology, Xinxiang Medical University, Xinxiang, China; MOE Key Laboratory for Neuroinformation, School of Life Science and Technology, University of Electronic Science and Technology of China, Chengdu, China; School of Psychology and Cognitive Science, East China Normal University, Shanghai, China; Department of Molecular Psychology, Institute of Psychology and Education, Ulm University, Ulm, Germany; Interchange Forum for Reflecting on Intelligent Systems, University of Stuttgart, Stuttgart, Germany; Brain Cognition and Brain Disease Institute (BCBDI), Shenzhen Institute of Advanced Technology, Chinese Academy of Sciences, Shenzhen, China; School of Psychological and Cognitive Sciences, Beijing Key Laboratory of Behavior and Mental Health, IDG/McGovern Institute for Brain Research, Peking. Tsinghua Center for Life Sciences, Peking University, Beijing, China

**Keywords:** serotonin, tryptophan, threat, anxiety, fear, depression, amygdala, PFC

## Abstract

Serotonin (5-HT) has long been implicated in adaptive emotion regulation as well as the development and treatment of emotional dysregulations in mental disorders. Accumulating evidence suggests that a genetic vulnerability may render some individuals at a greater risk for the detrimental effects of transient variations in 5-HT signaling. The present study aimed to investigate whether individual variations in the Tryptophan hydroxylase 2 (TPH2) genetics influence susceptibility for behavioral and neural threat reactivity dysregulations during transiently decreased 5-HT signaling. To this end, interactive effects between TPH2 (rs4570625) genotype and acute tryptophan depletion (ATD) on reactivity towards angry, neutral and happy faces were examined in a within-subject placebo-controlled pharmacological fMRI trial (n = 51). An a priori genotype stratification approach of extreme groups (GG vs. TT) allowed balanced sampling. While no main effects of ATD on neural reactivity to threat-related stimuli and mood state were observed in the entire sample, accounting for TPH2 genotype revealed an ATD-induced increase in subjective anxious arousal in the GG but not the TT carriers. The effects were mirrored on the neural level, such that ATD specifically reduced ventromedial prefrontal cortex (vmPFC) reactivity towards threat-related stimuli in the GG carriers. Furthermore, the ATD-induced increase in subjective anxiety positively associated with the extent of ATD-induced changes in vmPFC activity in response to threat-related stimuli in GG carriers. Together the present findings suggest for the first time that individual variations in TPH2 genetics render individuals susceptible to the anxiogenic and neural effects of a transient decrease in 5-HT signaling.

## Introduction

Serotonin (5-HT) has long been implicated in adaptive emotion regulation as well as the development and treatment of emotional dysregulations in mental disorders, in particular anxiety and mood disorders [1-4]. Most human research on the role of 5-HT in these domains is based on determining behavioral and neural variations associated with individual differences in central 5-HT signaling, including stable (genetic, trait-like) variations as well as transient variations e.g. induced by a rapid decrease in central serotonergic signaling via acute tryptophan depletion (ATD) procedures [5-7]. However, recent quantitative and qualitative reviews challenge both the role of 5-HT genetics in the etiology of emotional dysregulations in mood and anxiety disorders [4, 8] as well as the anxiogenic effects of ATD and associated neural processes [9-11].

Mounting evidence suggests considerable individual variations in the emotional effects of an ATD-induced transient reduction of central serotonergic signaling. Individual variations in the effects of ATD have been particularly described in the domains of fear/anxiety and executive control and linked to individual differences in 5- HT genetics and associated personality traits [12-18]. These findings may point to an underlying genetic vulnerability that renders some individuals at a greater risk for the detrimental effects of transient variations in 5-HT signaling. However, the few previous studies exclusively focused on genetic variations in the 5-HT transporter-linked promoter region (5-HTTLPR), a polymorphism with unclear relevance for both, central serotonergic signaling and psychopathological phenotypes [19, 20].

Central serotonergic signaling is directly regulated by the 5-HT biosynthesis rate limiting enzyme tryptophan hydroxylase 2 (TPH2) and variations in TPH2 genetics have been robustly linked to serotonin synthesis rates in limbic and prefrontal systems [21-23] and behavioral variations in emotional reactivity and regulation [24-26]. Initial studies employing genetic imaging approaches furthermore reported that TPH2 rs4570625 single nucleotide polymorphism (SNP) variants affect emotional reactivity in limbic regions and regulatory control in frontal regions [27-30]. The TPH2 rs4570625 SNP biases the reactivity of the amygdala in response to threatening stimuli [31] and attenuates top-down regulatory control of the ventromedial prefrontal cortex (vmPFC) [17]. Dysregulations in these regions may thus represent a candidate marker that neurally reflects a TPH-2 variation-related vulnerability for the detrimental effects of transient disruptions in 5-HT signaling.

Previous studies have revealed some - yet also inconsistent - evidence for effects of transiently reduced 5-HT signaling induced by ATD on the amygdala-vmPFC circuitry. ATD modulated communication in this circuitry and biased processing towards aversive emotional stimuli [32, 33]. ATD moreover affected the activity of the amygdala and the vmPFC during aversive stimuli processing [11, 34, 35], such that emotionally salient stimuli yielded higher responses in the mPFC as a result of impaired emotional evaluation following ATD [36, 37].

Despite accumulating evidence that interactions between genetic and pharmacological variations in 5-HT signaling affect the neural mechanisms of emotion processing, it remains unknown if individual variations the TPH2 gene polymorphisms can render individuals vulnerable for the effects of a transient variation in 5-HT signaling. The present study therefore examined whether the TPH2 SNP rs4570625 polymorphism determines the impact of an ATD-induced transient dysfunction in serotonergic signaling on subjective anxiety and on threat-specific neural reactivity of the vmPFC-amygdala circuitry (e.g. in responses to angry or fearful, but not happy faces) using a pre-registered within-subject randomized placebo-controlled double-blind fMRI experiment during which healthy male participants (n=51) either underwent transient decreases in 5-HT signaling (via a previously validated orally administered acute tryptophan depletion protocol, ATD) or a matched placebo protocol (PLC). Effects on threat-related amygdala-vmPFC reactivity were determined using functional magnetic resonance imaging (fMRI) during which angry faces and fearful faces (threat condition) as well as neutral, happy and non-emotional control stimuli were presented. First, we aimed to test whether the effect of diminished central 5-HT on threat-related amygdala-vmPFC reactivity varied as a function of TPH2 gene polymorphisms. Second, the relationship between ATD-induced effects on subjective anxious arousal and brain activity changes induced by ATD was explored. In particular, we hypothesized an interaction between the TPH2 genotype and ATD on subjective anxiety and amygdala-vmPFC activity, which together may reflect a genetic vulnerability factor for the development of emotional dysregulations in response to transient 5-HT changes.

## Method

### Participants

A total of 51 right-handed, healthy male participants (age *M* = 22.02 ± 2.48, 27 TT carriers, 24 GG carriers) were enrolled. Please note that TT and GG carriers were invited after a priori genotyping to increase statistical power for the present experiment. To reduce variance related to sex differences in 5-HT synthesis rates [38], only male subjects were included. In the context of problematic effect size determination in genetic imaging studies [39, 40] sample size was based on previous studies [41]. Individuals with a current or a history of medical, neurological/psychiatric disorders, regular use of psychotropic substances or MRI contraindications were excluded. The study was approved by the local ethics committee and in accordance with the latest revision of the Declaration of Helsinki. Written informed consent was obtained and the study was pre-registered on clinicaltrials.gov (https://clinicaltrials.gov/ct2/ show/NCT03549182, ID NCT03549182).

### Genotyping

Genomic DNA was purified from buccal cells using a MagNA Pure96 robot (Roche Diagnostics, Mannheim), sample probes were designed by TIB MolBiol (Berlin, Germany). Genotyping was conducted by real-time polymerase chain reaction (RT-PCR) and subsequent high-resolution melting on a Cobas Z480 Light Cycler (Roche Diagnostics, Mannheim, Germany) according to the manufacturer’s instructions.

### Serotonergic Challenge

Previous studies have demonstrated that after administration of ATD, tryptophan levels continuously decrease until they reach a plateau after about 5 hours and that the robust decrease lasts around two hours [6, 7, 42]. Given that tryptophan is the amino acid precursor of serotonin, the ATD procedure induces a transient and selective reduction in central serotonergic neurotransmission [5]. The contents of the amino-acid mixtures were based on previously validated proportions [43]. The amino acid mixture (ATD) consisted of 5.5g L-alanine, 4.9 g L-arginine, 2.7 g L-cystine, 3.2 g glycine, 3.2g L-histidine, 8g L-isoleucine, 13.5 g L-leucine, 8.9 g L-lysine, 3.0 g L-methionine, 12.2 g L-proline, 5.7 g L-phenylalanine, 6.9 g L-serine, 6.5 g L-threonine, 6.9 g L-tyrosine, 8.9 g L-valine, (total: 100 g). The control drink (PLC) contained identical ingredients plus 2.3 g of L-tryptophan (total: 102.3 g). The drinks were prepared by stirring the mixture into 200-ml water and lemon-lime flavor was added to mask the taste of the mixture.

### Control variables

To control for between-group differences (TT, GG) in anxiety, current stress, and early life stress, the State-Trait Anxiety Inventory (STAI) [44], Perceived Stress Scale (PSS) [45] and Childhood Trauma Questionnaire (CTQ) [46] were administered before treatment administration. To assess effects of treatment on subjective anxiety levels the State Anxiety Inventory component of the STAI (SAI) was repeatedly administered before administration of the amino acid drink (T1), during transient 5-HT decrease (immediately before MRI acquisition, T2) and at the end of the experiment (T3).

### Procedure

The present study employed a with-subject randomized double-blind placebo-controlled design. Participants were instructed to abstain from alcohol and caffeine for 24h and from food and drinks (except water) for 12h prior to the experiment. To adhere to the pharmacodynamic profile of treatment, participants arrived between 7:30 to 10:00 AM and underwent fMRI acquisition between 13:00 to 15:30 PM. Upon arrival, participants received a standardized protein-poor diet for breakfast. Following the assessment of pre-treatment control variables, participants underwent a previously validated tryptophan depletion protocol with ATD which has been demonstrated to lead to a robust transient reduction in central 5HT signaling [7, 43, 47, 48] or PLC. The ingestion of the amino acid mixtures was followed by a resting period of 5 hours to achieve a robust reduction in tryptophan levels. During the resting period participants were asked to relax and magazines were provided. Subsequently, participants underwent the fMRI paradigm. Control variables were re-assessed immediately before fMRI and after the emotion processing paradigms (at the end of the fMRI acquisition) (schematic outline of the experimental protocols see Figure 1). All subjects were administered both treatments (ATD, PLC) separated by approx. 5 weeks of wash-out period.

**Figure. 1.**
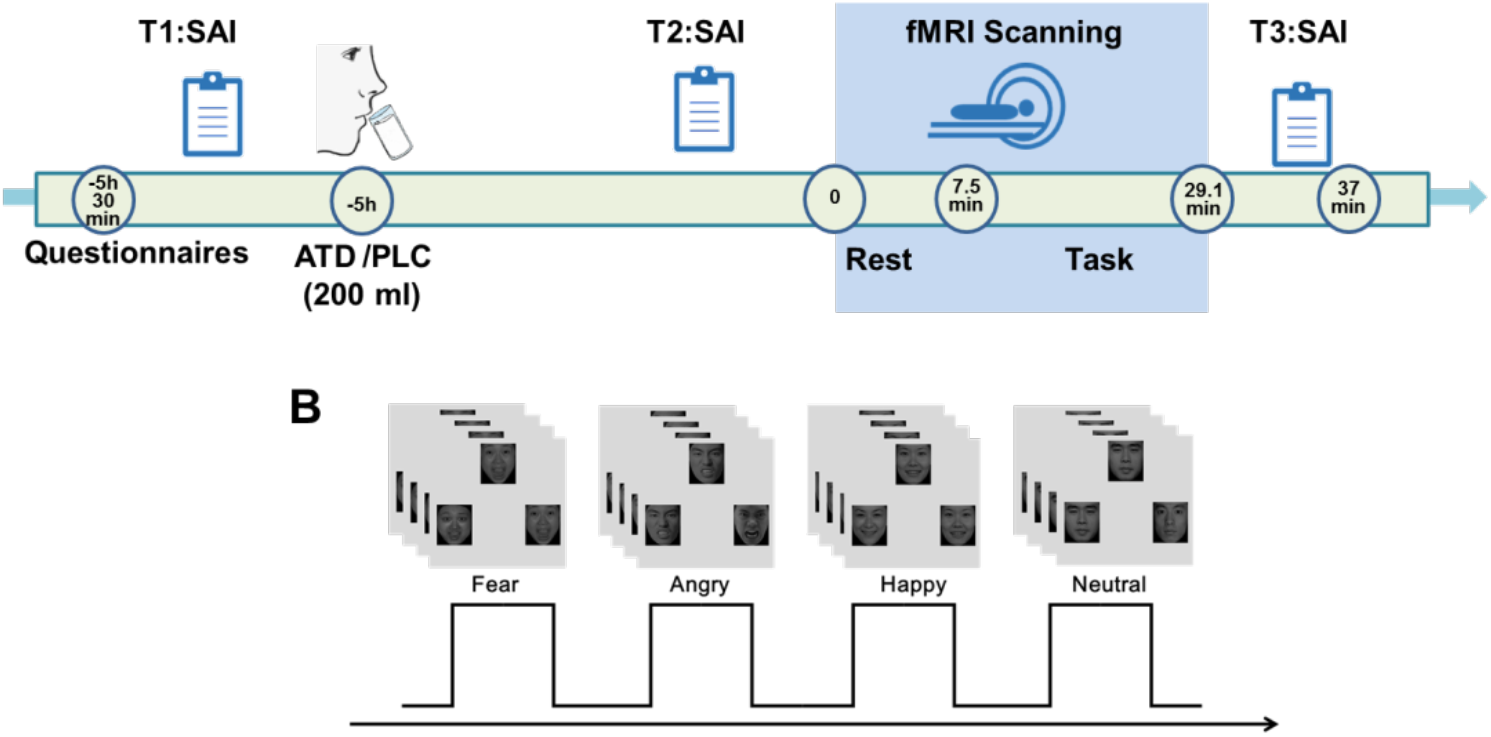
Experimental design and treatment protocols

### Experimental paradigm

A validated blocked-design emotional face-matching paradigm was administered during the two experimental sessions [49]. The paradigm consisted of 6 runs and every run comprised 4 blocks of facial stimuli as well as 1 block of non-facial stimuli serving as non-social control stimuli. During the face-processing blocks, a trio of condition-specific (angry, fearful, happy or neutral expressions) facial stimuli was presented and subjects were required to select one of the two faces (lower panel) that was identical to a target face (upper panel). Each block comprised four condition-specific trials, balanced for face gender. Asian facial stimuli were selected from a standardized Asian facial expression database [50]. During the non-social control blocks a trio of simple geometric shapes (circles and ellipses) was presented and subjects were required to select one of two shapes (lower panel) that matched the target shape presented in upper panel (Figure 1B). Each control block comprised four different shape trios. All blocks were preceded by a brief instruction (‘Face match’ or ‘Shape match’) that lasted 2s. Within each block, each trial was presented for 4s with a variable interstimulus interval (ISI) of 1-3 s (mean, 2s). The total block length was 24s and the total paradigm lasted 21min 36s.

### MRI data acquisition and processing

MRI data were acquired on a 3.0 Tesla GE MR750 system (General Electric Medical System, Milwaukee, WI, USA). To exclude subjects with apparent brain pathologies and improve normalization of the functional time series, T1-weighted high-resolution anatomical images were acquired with a spoiled gradient echo pulse sequence: repetition time (TR), 5.9ms; echo time (TE), minimum; flip angle, 9°; field of view (FOV), 256 × 256mm; acquisition matrix, 256 × 256; thickness, 1mm; number of slices, 156. For the functional MRI timeseries a total of 648 functional volumes were acquired using a T2*-weighted Echo Planar Imaging (EPI) sequence (TR, 2000ms; TE, 30ms; FOV, 220 × 220mm; flip angle, 90°; image matrix, 64 × 64; thickness/gap, 3.2/0mm; 43 axial slices with an interleaved ascending order).

Functional time-series were pre-processed using statistical parametric mapping (SPM12; Wellcome Department of Cognitive Neurology, Institute of Neurology, London, United Kingdom). For each subject and run the first seven volumes were discarded to allow magnet-steady images. The remaining functional images were slice- time corrected and realigned to the first image to correct for head motion. The functional images were next co-registered to the T1-weighted structural images and normalized to Montreal Neurological Institute (MNI) standard space using the segmentation parameters obtained from segmenting the structural images and interpolated at 3×3×3mm voxel size. Finally, the normalized images were spatially smoothed with an 8-mm full-width at half maximum (FWHM) Gaussian filter. For statistical analysis a two-step general linear model (GLM) approach was employed. At the single subject level an event-related general linear model (GLM) was employed including condition-specific regressors modelling the five experimental conditions, the instruction period and the six head motion parameters. Regressors for the experimental conditions and instruction were convolved with the default SPM hemodynamic response function (HRF). The design matrices additionally included a high pass filter to control for low frequency components and a first-order autoregressive model (AR[1]) to account for autocorrelation in the time-series.

### Region of Interest (ROI) selection and statistical thresholding

Based on our regional specific hypotheses derived from previous work [51-55], the analyses specifically focused on the amygdala and vmPFC. The amygdala ROIs were derived from Automatic Anatomical Labelling (AAL) [56]. The vmPFC ROI was defined based on an automated meta-analysis of 199 studies using NeuroSynth (www.neurosynth.org) with the search term “vmPFC” and thresholded at *p*_FDR_<0.05.

To balance analytical robustness and regional specificity we employed two analytic approaches: (1) To facilitate a robust determination of the effects an ROI analysis using extracted beta estimates from the vmPFC, left and right amygdala was employed. To determine emotion-specific interactions between genotype and treatment the extracted estimates were subjected to three-way ANOVAs (genotype×treatment×emotion) examining the effect for each ROI separately. The *p* values for the analysis were correspondingly adapted to *p* < 0.017 (0.05/3, corrected for three regions): (2) To facilitate a more specific localization of the effects, a voxel-wise small volume corrected analysis for the vmPFC and entire bilateral amygdala was additionally employed. To this end, a peak-level family-wise error (FWE) small volume correction was applied to a single mask encompassing the vmPFC and bilateral amygdala (total volume of the mask, 33912 mm^3^) (FWE, *p* < 0.05). Behavioral and neural (extracted parameter estimates) indices were further analyzed using SPSS 22.0 with appropriate ANOVA models and Bonferroni corrected post-hoc tests. *P* < 0.05 two-tailed was considered significant. Partial eta squared (*η*^*2*^_*p*_) and Cohen’s *d* were computed for as measures of effect size.

## Results

### Sample characteristics, confounders, genetic and treatment effects on subjective anxiety levels

The genotype groups did not differ with respect to pre-treatment control variables including age, trait anxiety, current stress, childhood trauma exposure and pre-treatment anxiety levels (all *p*s > 0.24, details see Table 1). Examining effects of treatment and genotype on state anxiety during the course of the experiment by means of mixed ANOVA models with genotype (TT vs. GG) as between-subject factor, and treatment (ATD vs. PLC) and timepoint (T1-T3; pre-oral administration, pre-fMRI, post-fMRI, T1-T3) as within-subject factors revealed a interaction effect between genotype and amino acid mixture (*F*_1,46_ = 4.67, *p* = 0.036, *η*^*2*^_*p*_ = 0.092), with post-hoc analyses suggesting that in GG carriers, participants reported higher anxious arousal levels during transient 5-HT decreases as compared to the PLC session (*p* = 0.013, Figure 2). Further exploratory analysis showed that the interaction effect was driven by a significant increase in state anxiety levels in the GG group during transient 5-HT decrease as compared to the PLC session following the threat exposure paradigm (T3; *t* = 2.83, *p* = 0.01). No other significant main and interaction effects were observed.

**Table 1.**
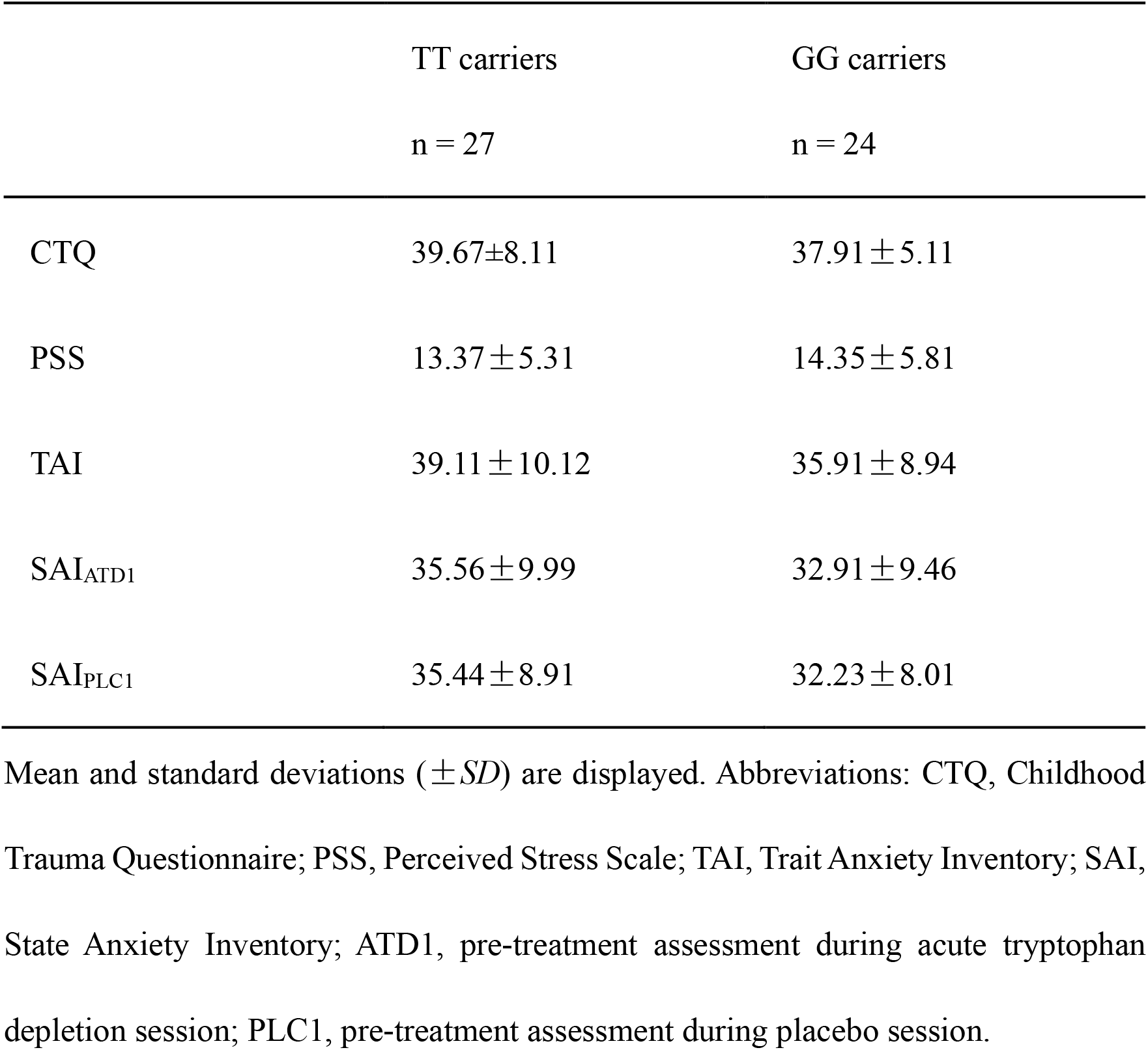
Sample characteristics (n = 51) stratified according to TPH2 rs4570625

**Figure. 2.**
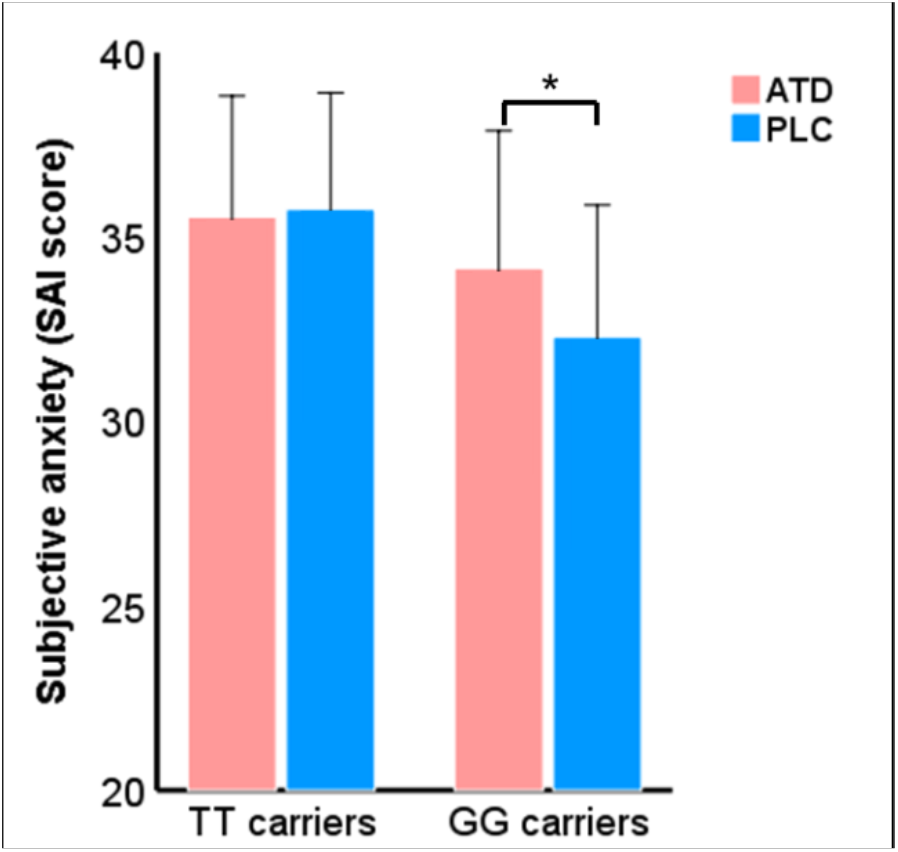
Results from the mixed ANOVA examining effects of treatment and genotype on anxious arousal levels over the course of the experiment. In GG carriers, participants reported higher state anxiety level in the ATD session as compared to the PLC sessions. ^*^*p* < 0.05, two-tailed.

### Behavioral results

Examining accuracy (ACC) and response times (RT) during the fMRI paradigm by means of mixed ANOVAs with emotion (angry vs. fearful vs. happy vs. neutral) and treatment (ATD vs. PLC) as within-subject factors and genotype (TT vs. GG) as between-subject factor revealed a significant main effect of emotion on both ACC (*F* = 15.93, *p* < 0.001, *η*^*2*^_*p*_ = 0.245) and RT (*F* = 115.58, *p* < 0.001, *η*^*2*^_*p*_ = 0.702). For the ACC, post-hoc analysis revealed that, all participants responded more accurately for angry (0.984±0.004) and fearful faces (0.980±0.003) as compared to happy (0.959±0.006) and neutral faces (0.956±0.007, all *p*s < 0.001), reflecting attentive task engagement irrespective of treatment. In addition, post-hoc analyses revealed that RTs for fearful face (1038.24 ± 23.77) were faster compared to angry faces (1120.72 ± 26.57), angry faces compared to happy faces (1186.84 ± 34.23) and slower RT for neutral faces (1272.39 ± 33.75) compared to all other conditions (all *p*s < 0.001). Importantly, we observed a significant main effect of treatment (*F* = 5.81, *p* = 0.02, *η*^*2*^_*p*_ = 0.106) suggesting that participants made generally slower responses in the ATD session as compared to PLC sessions (*p* = 0.02).

### Neural activity: ROI analysis - three-way interaction in vmPFC and amygdala to face emotions

Examination of the extracted parameter estimates from the vmPFC, left and right amygdala revealed a significant three-way interaction in the ROI-specific ANOVAs for both the vmPFC (*F*_3,147_ = 4.58; *p* = 0.007; *η*^*2*^_*p*_ = 0.085; power (1 -*β*) = 0.838) and right amygdala (*F*_3,147_ = 4.40; *p* = 0.006; *η*^*2*^_*p*_ = 0.082; power (1 - *β*) = 0.85), but not the left amygdala (*p* = 0.087). Post-hoc analyses on the extracted beta values from the vmPFC mask revealed that in the GG carriers, ATD treatment decreased vmPFC reactivity to fearful and angry face relative to the corresponding conditions under PLC treatment (fearful: *p* = 0.034, angry: *p* = 0.043), while for the TT carriers, ATD treatment suppressed vmPFC reactivity to happy face relative to the PLC treatment. Post-hoc analyses on the extracted beta values from the right amygdala mask revealed that during the PLC sessions, TT carriers showed higher reactivity both to happy face (*p* = 0.039) and neutral face (*p* = 0.037) as compared to GG carriers, but when participants received ATD treatment, there was no significant difference between GG and TT carriers (see Figure 3).

**Figure. 3.**
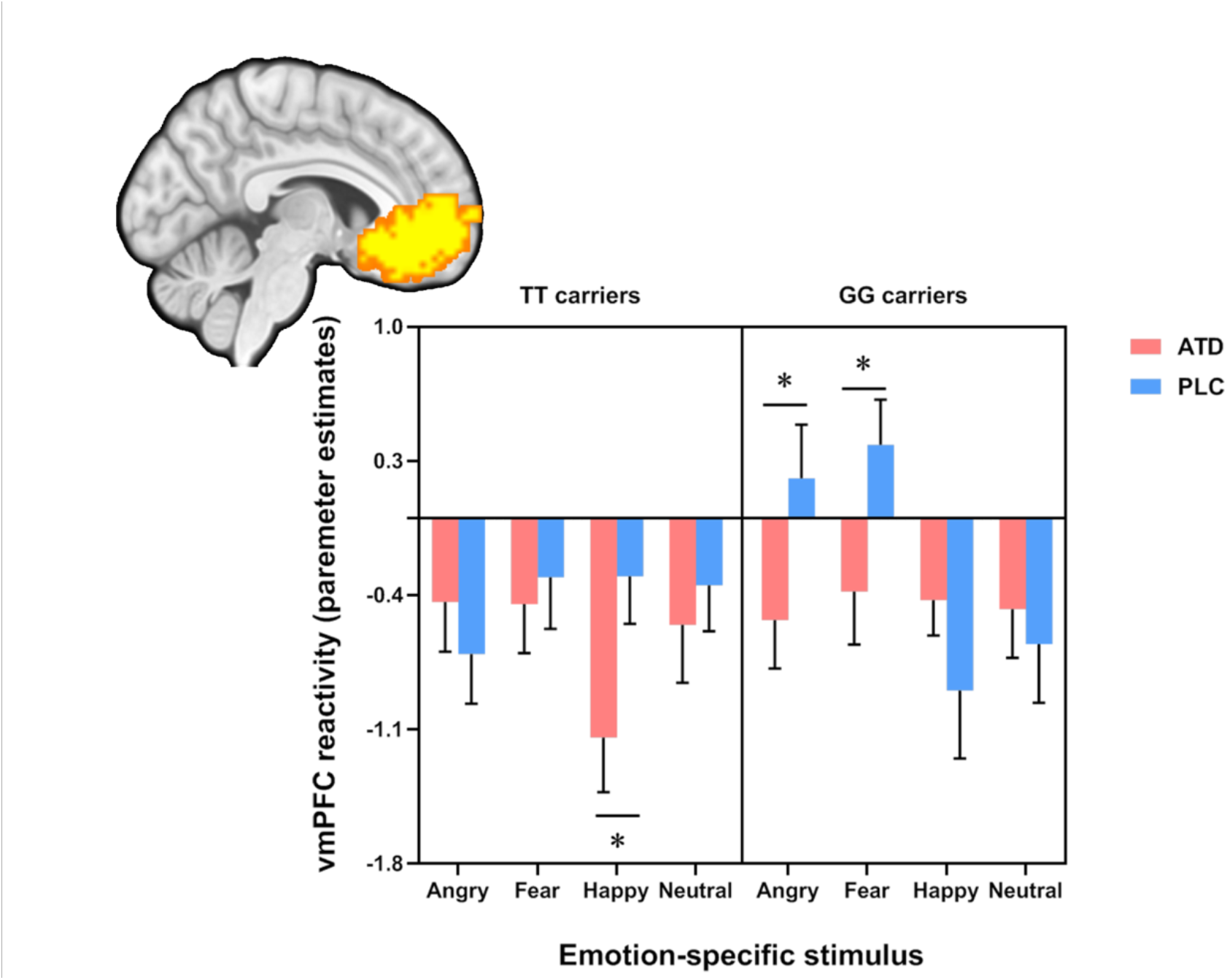
Results from a three-way ANOVA (genotype×treatment×emotion) examining the effect on vmPFC activation (defined by a NeuroSynth meta-analysis, shown in the upper left corner). Post-hoc analyses on the extracted beta values from the vmPFC mask revealed that in the GG carriers, ATD treatment decreased vmPFC reactivity to fearful and angry face relative to both the PLC treatment, while for the TT carriers, ATD treatment suppressed vmPFC reactivity to happy face relative to the PLC treatment. ^*^*p* < 0.05, two-tailed.

To facilitate a more specific localization of the effects, a voxel-wise small volume corrected analysis for the vmPFC and entire bilateral amygdala was additionally employed. This analysis revealed a significant three-way interaction effect in the vmPFC after small volume correction for the mask encompassing the vmPFC and entire amygdala (k = 103, *p*_FWE_ = 0.031, *F*_3, 343_ = 7.47, peak MNI co-ordinate: x = -6, y = 27, z = −6) (Fig. 4a), while no significant interaction effect was found in the right amygdala (*p*_FWE_ = 0.200). Thus, the interaction effect between genotype, treatment and emotion in the vmPFC appears to be robust, while interaction in right amygdala was less robust.

**Figure. 4.**
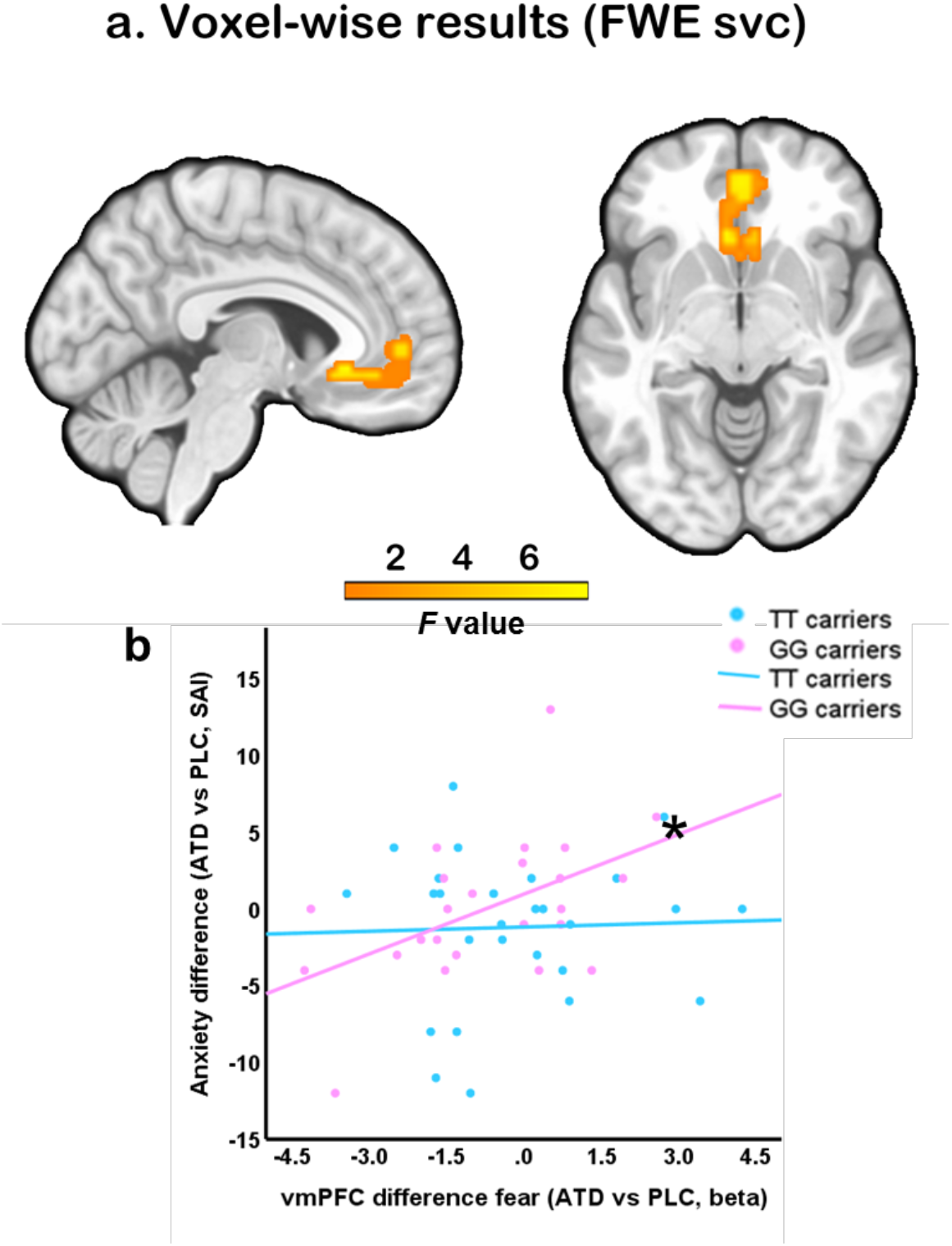
Regional-specific analysis of the effects and associations between treatment-induced anxiety increased and treatment-induced alterations in the vmPFC towards fearful faces. (a) Precise localization of the coordinates in the vmPFC as determined by a voxel-wise ANOVA in combination with a small-volume correction applied to an anatomical mask covering the entire amygdala-vmPFC system. (b) Associations between anxiety level changes induced by treatment and brain response to fearful face changes induced by treatment. Abbreviations: anxiety difference (ATD vs PLC, SAI): SAI scores_ATD_ – SAI scores_PLC_; vmPFC difference fear (ATD vs PLC, beta): parameter estimates from the vmPFC mask for the contrast: fearful face_ATD_ – fearful face_PLC_, ^*^*p* < 0.05, two-tailed.

### Associations between subjective anxious arousal and neural activity

To explore the relationship between treatment-induced changes on the level of subjective experience and neural activity (T2: pre-fMRI) an exploratory correlation analysis was conducted. Results indicated a significant positive correlation between state anxiety level changes and vmPFC activity changes induced by treatment (*r* = 0.502, *p* = 0.013; Figure 4b) in GG carriers, while not in TT carriers (*r* = 0.035, *p* = 0.860, between group correlation difference *t* = 1.85, *p* = 0.065). No significant associations were observed for the other face conditions (all *p*s>0.05).

## Discussion

The present study aimed at determining whether individual differences in a genetic polymorphism with a regulatory role over central serotonin synthesis rates (TPH2) influence susceptibility to the effects of a transient decrease in 5-HT signaling on subjective anxiety and threat-related neural activity. To this end interactions between TPH2 genotype and the effects of ATD were examined in a pre-registered within-subject placebo-controlled pharmacological fMRI trial. Partly resembling previous findings, the present study did not reveal significant effects of ATD on subjective anxiety or neural reactivity to threat-related stimuli in the entire (pooled) sample [10, 57, 58]. However, when accounting for genetic differences significant interaction effects between TPH2 genotype and ATD were observed, such that specifically the GG carriers reported increased anxious arousal after ATD, whereas TT carriers reported no significant changes during transiently decreased 5-HT signaling. The genotype-specific anxiogenic effects were mirrored on the neural level, such that ATD specifically reduced vmPFC reactivity towards threat-related stimuli in the GG carriers. In contrast, in TT carriers ATD – relative to PLC – induced a reduced reactivity to happy faces in the vmPFC. Finally, in GG carriers the ATD-induced increase in subjective anxiety was positively associated with the extent of ATD-induced changes in vmPFC activity in response to fearful faces following ATD. Together the present findings demonstrate for the first time that individual variations in TPH2 genetics render healthy participants susceptible to the anxiogenic and neural effects of a transient decrease in 5-HT signaling.

The relevance of the serotonergic system for anxiety-related processes has long been proposed and has been supported by several findings [59-62]. Several of these studies have capitalized on a validated ATD procedure which allows a reliable and transient decrease in central serotonin synthesis rates. While ATD reliably decreases the availability of tryptophan - the precursor of serotonin - its anxiogenic effects have been recently questioned in a comprehensive meta-analysis [10] and accumulating evidence suggests strong individual differences [12-14, 16, 18, 63]. In line with our hypothesis for a genetically determined susceptibility to the subjective emotional effects of ATD we found that specifically GG carriers experienced anxiogenic effects. Although the neurobiological implications of the TPH2 rs4570625 polymorphism are not fully understood, the G-allele has been suggested to be related to a hypofunction of tryptophan hydroxylase [64, 65] and in turn lower 5-HT synthesis rates which may promote a stronger impact of a transient decrease in tryptophan.

Consistent with our hypothesis, the susceptibility on the behavioral level was paralleled by a threat-specific interaction effect in the vmPFC-amygdala circuits. Specifically in GG carriers ATD induced a decreased vmPFC-response to threat-related stimuli, i.e. fearful and angry faces. Animal model and human studies demonstrated that serotonergic fibers project into the limbic system and cortical regions, including the vmPFC [52, 66], suggesting that 5-HT regulates neural activity in these circuits.

In the context of anxiety-related domains the vmPFC plays a critical role in both motivational processes to minimize threat [67-69] and implicit emotion regulation of threat, i.e. during extinction [53, 70]. Previous studies in individuals with pathological anxiety have reported deficient engagement of the vmPFC, including decreased vmPFC engagement accompanied by deficient differentiation of safety from threat [71] in generalized anxiety disorder or deficient extinction recall across anxiety disorders [72]. The associated deficits in safety-threat differentiation may in turn bias both the perception of information as threatening [73] and an adaptive regulatory control over threat. In line with this proposed role of the vmPFC, non-invasive stimulation of this region attenuates the attentional bias to threat-related stimuli in the dot-probe test [67] and facilitates adaptive threat extinction [74]. The observation of ATD-induced decreased vmPFC reactivity to threat-related stimuli associated with the extent of treatment-induced increased anxiety levels in the GG carriers may thus reflect either an enhanced detection of threat or a reduced engagement of implicit top-down fear regulation, both of which have been associated with higher anxious arousal [53, 75].

In contrast to our hypothesis, no robust treatment and genotype interaction effects were observed on the mean amplitude of amygdala threat reactivity. These findings may underscore that the genetic vulnerability to ATD renders prefrontal regulatory regions but not the amygdala - which is more involved in bottom-up threat reactivity – susceptible to transient serotonergic hypoactivity. Moreover, the amygdala shows a rapid adaptation to repeated presentations with both reduced [76-78] and increased reactivity [54, 79-81] being reported, which may have overshadowed determination of robust effect. In addition, previous evidence between amygdala reactivity and 5-HT functioning is inconsistent, withsome studies failing to find associations between general amygdala reactivity and 5-HT genetics as well as robust effects of ATD on amygdala activity [9, 82, 83].

Given the inconsistent results on associations between serotonergic functioning and disorders characterized by emotional regulation deficits the present findings have additional clinical implications. First, transient changes in tryptophan and serotonin levels have been associated with different environmental factors such as psychosocial stress, inflammation, and dietary habits [84], and the present results indicate that particular individuals with a specific TPH2 genotype are vulnerable to the detrimental effects on anxiety and neural activity. The underlying changes may represent a core pathological mechanism for the development and maintenance of exaggerated anxious arousal which can promote emotional dysregulations and mental disorders. Second, serotonin dysfunction has been suggested as a candidate biomarker for exaggerated anxiety [85] and promising treatment target for anxiety disorders. The present results indicate that effects of serotonin-modulation on anxiety and associated vmPFC threat responses depend on individual variation in the TPH2 gene. This can lead to individual variations in the initial response to treatments targeting serotonin transmission, including selective serotonin reuptake inhibitors (SSRIs) frequently prescribed for anxiety and mood disorders.

Findings of the present study need to be considered in the context of the following limitations. First, only male subjects were enrolled to reduce variance in the data related to sex differences in 5-HT synthesis [38]. Future studies need to determine whether the observed effects generalize to women. Second, considering that drawing blood samples might increase anxiety levels and thus can confound our primary outcomes tryptophan blood levels were not assessed. However, previous studies reported robust and selective decreases in 5-HT signaling following similar the ATD treatment protocols as used in the present study [43, 86], the additional examination of blood-level measures particularly in the combined treatment group may reveal important additional information on the relevance of serotonergic system to anxiety-relevant processes and should be included in future studies. The present study matched the groups for trait anxiety, while a number of studies reported associations with trait anxiety impact cognitive and emotional processes as well as pharmacological effects [58, 87, 88]. Finally, while the study involved a total of 51 participants in a complex pharmaco-fMRI design we did not include an a priori sample size calculation. The priori calculation for sample size and power in complex genetics-pharmaco-fMRI design is still limited although our sample size was based on previous studies [41]. Although the ROI analysis may have facilitated a more robust determination of effects validation and replication designs are needed [89, see also 23].

Together, the present findings provide the first evidence that individual variations in a TPH2 polymorphism – a genetic variation functionally related to central 5-HT signaling – render individuals susceptible for the anxiogenic and threat-specific neural effects of transient variations in serotonergic signaling. They may also explain the previous inconsistent findings on the effects of acute tryptophan depletion in healthy individuals and point to a vulnerability marker for conditions characterized by excessive anxiety.

## Data Availability

Unthresholded group-level statistical maps are available on NeuroVault (https://neurovault.org/images/794826/), additional data related to study is available from the corresponding author upon reasonable request

## Acknowledgements

This work was supported by the China MOST2030 Brain Project (Grant No. 2022ZD0208500), National Key Research and Development Program of China (Grant No. 2018YFA0701400) and the National Natural Science Foundation of China (Grants No. 32250610208, 82271583)

## Author contributions

CL and BB designed the study. CL, KL, XZ, MF, CS, YZ conducted the experiment and collected the data. CL performed data analyses. CL and BB wrote the manuscript. HZ, YS, BZ, CM, ZW and KK revised the manuscript draft.

## Data availability

Unthresholded group-level statistical maps are available on NeuroVault (https://neurovault.org/images/794826/), additional data related to study is available from the corresponding author upon reasonable request. A preprint of the manuscript had been archived on the biorxiv.org repository.

